# Divergent patterns of cognitive decline in preclinical Alzheimer’s disease: implications for secondary prevention trials

**DOI:** 10.64898/2025.12.15.25342305

**Authors:** Runpeng Li, Oliver Langford, Philip S. Insel, Reisa A. Sperling, Rema Raman, Paul S. Aisen, Michael C. Donohue

## Abstract

**Background:** Alzheimer’s disease biomarkers in cognitively unimpaired older adults are associated with later cognitive and clinical decline, yet substantial heterogeneity in the timing and rate of decline remains insufficiently characterized. This study aims to identify subgroups of cognitive decline among biomarker-defined cognitively unimpaired adults and determine baseline predictors of heterogeneity in preclinical Alzheimer’s disease progression.

**Methods:** Longitudinal data was analyzed from the Anti-Amyloid Treatment in Asymptomatic Alzheimer’s Disease (A4) Study, which enrolled amyloid-positive participants, and the parallel LEARN Study, which enrolled amyloid-negative individuals meeting all other A4 criteria. Participants completed baseline amyloid PET, plasma P-tau217, structural MRI, and serial cognitive assessments. Latent Class Mixed-Effects Models (LCMMs) were used to identify distinct cognitive trajectory classes. Associations between class membership and demographic, clinical, and biomarker characteristics were evaluated. The primary outcome was longitudinal change in the Preclinical Alzheimer Cognitive Composite (PACC).

**Findings:** Three cognitive trajectory classes were identified: stable, slow decliners, and fast decliners. Higher plasma P-tau217, smaller hippocampal volume, and elevated tau PET were associated with greater odds of belonging to declining classes. Among amyloid-positive individuals, approximately 70% were classified as stable over the observed follow-up interval. These stable individuals likely contribute little to the power of preclincal Alzheimer’s trials.

**Interpretation:** Latent class modeling reveals marked heterogeneity in preclinical cognitive trajectories, even among individuals with biomarker evidence of Alzheimer pathology. The high proportion of stable individuals, though consistent with the long presymptomatic interval, has important implications for prevention trial design, particularly regarding inclusion criteria, outcome measures, and treatment effect assumptions. Identifying subgroups of decline may improve prognostic modeling and guide enrichment strategies for precision secondary prevention trials.

**Funding:** US National Institutes of Health, Eli Lilly, Alzheimer’s Association, Foundation for the National Institutes of Health, GHR Foundation, Davis Alzheimer Prevention Program, Yugilbar Foundation, Avid Radiopharmaceuticals, Cogstate, Albert Einstein College of Medicine, Foundation for Neurologic Diseases, and Epstein Family Foundation.

**Research in Context:** *Evidence before this study:* We searched PubMed, MEDLINE, and Google Scholar for articles published between January 1, 2000, and December 31, 2025, using the terms “preclinical Alzheimer’s disease,” “cognitive decline,” and “latent class.” Previous studies have identified patterns of cognitive decline in aging populations, but there is limited understanding of the predictive value of recently developed biomarkers of Alzheimer’s pathology, and we found no studies examining the impact on secondary prevention trial design.

*Added value of this study:* This study is the first to apply latent class analysis to data from the A4 and LEARN studies, identifying distinct classes of cognitive decline in individuals at risk for Alzheimer’s disease. Our findings reveal significant heterogeneity in cognitive trajectories, suggesting that preclinical Alzheimer’s disease is not a uniform process. These insights could inform personalized intervention strategies and improve the design of future clinical trials.

*Implications of all the available evidence:* Our results underscore the importance of considering individual variability in cognitive decline when developing and testing interventions for preclinical Alzheimer’s disease. Recognizing the diverse patterns of cognitive decline can enhance the precision of clinical trial designs and lead to more effective, tailored therapeutic approaches. Future research should focus on validating these findings in larger, more diverse cohorts and exploring the underlying mechanisms driving the observed heterogeneity.

## Introduction

Elevated levels of brain amyloid in cognitively unimpaired older adults are associated with subsequent cognitive decline and increased risk of clinical progression.^1–5^ The Anti-Amyloid Treatment in Asymptomatic Alzheimer’s Disease (A4) Study, a randomized trial of solanezumab in cognitively normal individuals with elevated amyloid PET, demonstrated group-level decline on a cognitive composite but no treatment benefit.^2^ Follow-up analyses from A4 and its companion observational study of amyloid-negative individuals, the Longitudinal Evaluation of Amyloid Risk and Neurodegeneration (LEARN), further confirmed that higher baseline amyloid PET and plasma phosphorylated tau (P-tau217) levels are associated with faster cognitive decline and increased functional progression.^5^

Despite these consistent group-level associations, cognitive trajectories among amyloid-positive individuals remain highly variable. Conventional longitudinal models assume that individual cognitive trajectories are randomly scattered about a single mean trend for given covariate values, potentially obscuring meaningful heterogeneity. To better characterize this heterogeneity, we applied a Latent Class Mixed-Effects Model (LCMM)^6^ to A4 and LEARN data. This approach identifies unobserved subgroups of participants with distinct longitudinal patterns of change and allows for evaluation of baseline biomarkers and demographic factors as predictors of class membership.

Latent class growth model approaches have been applied several times in the literature to identify distinct classes of late-life cognitive trajectories. A systematic review by Wu et al.^7^ identified 37 investigations and these consistently identified three classes: stable, slow decline, and fast decline. However, many of these papers lacked more recently developed biomarkers of Alzheimer’s pathology. One exception is the study by Teipel et al.^8^ included amyloid PET as a predictor in a cohort of *N* = 265 individuals followed for two years. Villeneuve et al.^9^ identified latent classes of Amsterdam Instrumental-Activities-of-Daily-Living questionnaire using amyloid PET and FDG PET in N=289 participants over three years. Here, we consider latent classes of cognition predicted by APOE genotype, amyloid PET, P-tau217, hippocampal atrophy, and tau PET in a cohort of *N* = 1, 629 individuals followed for up to seven years in A4 and LEARN. Furthermore, we explore practical implications for classification algorithms and clinical trials.

## Methods

### Conduct of the A4 and LEARN Studies

The A4 and LEARN studies have been previously described^2,5^, but we briefly summarize key elements. The A4 Study was a multicenter, randomized, double-blind, placebo-controlled secondary prevention trial enrolling cognitively unimpaired older adults aged 65–85 years with elevated brain amyloid on florbetapir PET. Participants were recruited from 67 sites in the United States, Canada, Australia, and Japan and underwent standardized screening, baseline clinical assessment, biomarker acquisition, and neuropsychological testing. After eligibility confirmation, participants were randomized to receive solanezumab or placebo and were assessed longitudinally with regular cognitive evaluations, safety monitoring, and follow-up imaging and fluid biomarker collection per protocol.

The LEARN study was conducted in parallel as an observational cohort enrolling individuals who met all A4 screening criteria except for amyloid PET positivity. LEARN participants completed identical clinical and cognitive assessments, allowing direct comparison of trajectories between amyloid-positive and amyloid-negative individuals. Both studies followed harmonized procedures for data collection, visit scheduling, and quality control to ensure comparability of longitudinal outcomes.

All participants of both studies provided written informed consent, and study protocols were approved by the institutional review boards or ethics committees at each participating site.

### Statistical Analysis

#### Latent Class Mixed-Effects Models

Latent class mixed-effects models (LCMMs) extend traditional mixed-effects models by identifying unobserved subgroups, or latent classes, of individuals who follow distinct longitudinal trajectories. In the context of cognitive decline in Alzheimer’s disease, LCMMs allow for the estimation of population-level trends while capturing individual variability and uncovering hidden subpopulations that may progress at different rates.

Analyses were conducted using harmonized data from the A4 and LEARN studies, accessed through the A4LEARN R data package (version 1.1.20250808^10,11^). This package provides curated datasets and metadata derived from the A4 and LEARN clinical studies for reproducible statistical analysis. Code to reproduce the results from this paper are available from the A4LEARN GitHub repository^12^.

The dependent variable was the Box-Cox-transformed Preclinical Alzheimer Cognitive Composite (PACC)^13^ score at each visit. The Box-Cox transformation parameter (*λ*) was selected to maximize the likelihood that the transformed data approximated normality. Time was modeled as a continuous variable representing years since baseline.

Natural cubic spline basis functions with one, two or three degrees of freedom were evaluated to model nonlinear trajectories, with boundary knots at baseline and the maximum follow-up time and an interior knots at the median or tertiles of observation time. The choice of degrees of freedom and the number of classes (also one, two, or three) was determined by model fit criteria (Bayesian Information Criterion [BIC] and Integrated Complete Likelihood [ICL]).^14^ Each model included class-specific spline-based time effects, as well as subject-specific random intercepts. The effect of baseline covariates were shared across latent classes and included: randomized to solanezumab (1 for the active group, 0 for placebo or LEARN), plasma P-tau217 concentration, florbetapir cortical standardized uptake value ratio (SUVr), APOE *∈*4 carrier status, sex, age, education, hippocampal atrophy, and PACC test stimulus version administered (which alternated per study protocol). Plasma P-tau217 concentrations were measured using an immunoassay developed by Eli Lilly and reported in arbitrary units per milliliter (U/mL), where “U” denotes a generic assay unit proportional to signal intensity. Absolute calibration against mass concentration (e.g., pg/mL) was not available. Hippocampal atrophy measures were derived by first residualizing with respect to intracranial volume, and then standardizing to mean zero and variance one (i.e. z-scoring).

The LCMM includes two linked submodels. The longitudinal submodel captures within-class trajectories of cognitive change over time. Each class had its own set of spline coefficients, allowing class-specific shapes of decline. The class membership submodel defines the probability of belonging to each latent class as a multinomial logistic function of baseline covariates (which includes all covariates listed above except spline terms and PACC test version).

In a secondary analysis, we fit the model to the subset of participants with baseline flortaucipir (tau) PET imaging. Tau PET was summarized as the mean SUVr across eight cortical regions: entorhinal cortex, inferior temporal, inferior parietal, posterior cingulate, caudal middle frontal, middle temporal, superior parietal, and frontal pole. This composite measure was included as an additional covariate in both the longitudinal and class membership submodels to assess whether tau pathology improved prediction of cognitive decline patterns or latent class assignment.

To assess the reliability and potential predictive utility of the latent class membership model, we examined the posterior probabilities of class assignment for each participant. High posterior probabilities indicate greater certainty in latent class classification. We summarized these distributions across classes and computed the mean posterior probability within each group as an index of classification confidence.

#### Power Analysis for Latent Class-Specific Trials

To explore the impact of heterogeneity on clinical trials in preclinical Alzheimer’s disease, we conducted power calculations to estimate power for hypothetical trials targeting latent classes of cognitive decline among amyloid-positive participants. Effect size estimates were derived from longitudinal models using natural cubic splines applied to Preclinical Alzheimer Cognitive Composite (PACC) scores.^15^ Separate models were fit for (a) the stable class, (b) the decliner classes, and (c) a reference group of amyloid-negative stable individuals. The models included fixed effects for time (two degrees of freedom), APOE *ε*4 carrier status, sex, age, education, plasma P-tau217, and florbetapir PET. Residuals were assumed to be normally distributed with an unstructured covariance matrix.

For each group, we extracted mean PACC and residual variance from the model at two time points: 2 years and 4 years, representing potential trial durations. These estimates were used to compute the power to detect treatment effects. Power was approximated using two-sample t-test calculations, assuming 500 participants per arm, attrition rates of 10% at 2 years and 20% at 4 years, and a two-sided alpha of 0.05.

#### Ten-fold cross-validation of latent class predictions

To further evaluate prospective discrimination, we performed ten-fold cross-validation stratified by P-tau217 and latent class. For each of the ten folds, 90% of the data were used to re-train the LCMM, and the retrained model was used to predict latent classes for the 10% held-out test set using only baseline data. Model performance was summarized using the area under the precision-recall curve (AUPRC) for discrimination of each class versus all others. This approach can be more informative than receiver operating characteristic (ROC) curves when group sizes are imbalance.^16^

#### Regression Tree Analysis for Latent Class Characterization

To evaluate whether latent classes could be distinguished by baseline characteristics through a combination of binary decision rules, we conducted a post-hoc classification tree analysis^17,18^. Class assignments from the optimal latent class mixed model were used as the outcome variable, with baseline demographic and clinical characteristics as predictors (randomized to solanezumab, plasma P-tau217 concentration, florbetapir PET, APOE *ε*4 carrier status, sex, age, education, hippocampal atrophy, and PACC). We separately considered a regression tree with tau PET as an additional predictor.

Hyperparameter tuning was performed using 10-fold cross-validation. The optimal hyperparameters were selected based on maximum cross-validated accuracy. The final tree structure was visualized to illustrate the hierarchy of decision rules for class assignment. Variable importance was calculated based on the total reduction in node impurity attributed to splits on each predictor. This approach provides an interpretable framework for understanding the multivariate profile of each latent class and assessing whether classes can be reliably separated using clinically available baseline information.

All analyses were conducted using R (version 4.5.2; R Foundation for Statistical Computing) with the lcmm package (version 2.2.1).

## Results

### Study Participants

A total of *N* = 1, 629 participants were included in the analytic sample for the base model (without tau PET), comprising *N* = 1, 110 from the A4 Study and *N* = 519 from the LEARN study. The mean (standard deviation [SD]) age at baseline was 71.46 (4.69) years, and 60.2% were female. The median follow-up time was 6.0 years (interquartile range 3.9 to 7.0). Baseline characteristics stratified by latent class membership are presented in Table 1. A total of *N* = 427 individuals had tau PET data and were submitted to the tau PET model. The tau PET subset included *N* = 372 from the A4 Study and *N* = 55 from the LEARN study. Baseline characteristics for the tau PET subset are presented in Supplementary Table S1.

**Table 1:** Baseline Demographic, Clinical, and Biomarker Characteristics by Latent Class of Cognitive Decline. Baseline characteristics of participants classified by the latent class mixed model (LCMM) of Preclinical Alzheimer Cognitive Composite (PACC) trajectories. Classes represent distinct longitudinal cognitive patterns: stable, slow decliner, and fast decliner. Continuous variables are presented as mean (SD); categorical variables as No. (%). **Abbreviations:** APOE = apolipoprotein E; PET = positron emission tomography; P-tau217 = plasma phosphorylated tau 217; SUVr = standardized uptake value ratio; U/mL = arbitrary units proportional to assay signal; Am. = American; PI = Pacific Islander; CDR = Clinical Dementia Rating; Prog. = Progressor. **Footnotes:** Amyloid PET and tau PET SUVr values represent mean cortical uptake relative to cerebellar reference region. Hippocampal atrophy are residualized for intracranial volume and z-scored. Education reported in years of formal schooling. CDR Progressors were observed to have a CDR Global score greater than zero at two consecutive visits, or their last visit.

|  | Stable<br>(N=1257) | Slow decliner<br>(N=253) | Fast decliner<br>(N=119) | Total<br>(N=1629) | p value |
| --- | --- | --- | --- | --- | --- |
| Group |  |  |  |  | < 0.001 |
| - LEARN | 481 (38.3%) | 35 (13.8%) | 3 (2.5%) | 519 (31.9%) |  |
| - Placebo | 386 (30.7%) | 128 (50.6%) | 49 (41.2%) | 563 (34.6%) |  |
| - Solanezumab | 390 (31.0%) | 90 (35.6%) | 67 (56.3%) | 547 (33.6%) |  |
| Age, mean (SD), y | 70.78 (4.29) | 74.06 (5.24) | 73.10 (5.12) | 71.46 (4.69) | < 0.001 |
| Sex, No. (%) |  |  |  |  | 0.016 |
| - Male | 494 (39.3%) | 117 (46.2%) | 37 (31.1%) | 648 (39.8%) |  |
| - Female | 763 (60.7%) | 136 (53.8%) | 82 (68.9%) | 981 (60.2%) |  |
| Race, No. (%) |  |  |  |  | 0.773 |
| - N missing | 10 | 2 | 1 | 13 |  |
| - Am. Indian or Alaska Native | 6 (0.5%) | 0 (0.0%) | 1 (0.8%) | 7 (0.4%) |  |
| - Asian | 25 (2.0%) | 4 (1.6%) | 4 (3.4%) | 33 (2.0%) |  |
| - Native Hawaiian or Other PI | 0 (0.0%) | 0 (0.0%) | 0 (0.0%) | 0 (0.0%) |  |
| - Black or African Am. | 30 (2.4%) | 6 (2.4%) | 1 (0.8%) | 37 (2.3%) |  |
| - White | 1179 (94.5%) | 240 (95.6%) | 112 (94.9%) | 1531 (94.7%) |  |
| - Unknown or Not Reported | 7 (0.6%) | 1 (0.4%) | 0 (0.0%) | 8 (0.5%) |  |
| Ethnicity, No. (%) |  |  |  |  | 0.535 |
| - Hispanic or Latino | 35 (2.8%) | 8 (3.2%) | 5 (4.2%) | 48 (2.9%) |  |
| - Not Hispanic or Latino | 1211 (96.3%) | 241 (95.3%) | 114 (95.8%) | 1566 (96.1%) |  |
| - Unknown or Not reported | 11 (0.9%) | 4 (1.6%) | 0 (0.0%) | 15 (0.9%) |  |
| Education, mean (SD), y | 16.62 (2.74) | 16.58 (2.70) | 16.32 (2.85) | 16.59 (2.74) | 0.512 |
| APOE 4, No. (%) |  |  |  |  | < 0.001 |
| - non-carrier | 732 (58.2%) | 93 (36.8%) | 31 (26.1%) | 856 (52.5%) |  |
| - carrier | 525 (41.8%) | 160 (63.2%) | 88 (73.9%) | 773 (47.5%) |  |
| PACC, mean (SD) | 0.74 (2.37) | -0.90 (2.47) | -2.23 (3.10) | 0.27 (2.61) | < 0.001 |
| Amyloid PET, mean (SD), CL | 37.40 (34.81) | 72.10 (42.88) | 88.62 (37.75) | 46.53 (40.23) | < 0.001 |
| P-tau217, mean (SD), U/ml | 0.20 (0.09) | 0.33 (0.16) | 0.45 (0.26) | 0.24 (0.15) | < 0.001 |
| Hipp. atrophy, mean (SD), z-score | -0.20 (0.96) | 0.60 (0.96) | 0.95 (0.87) | 0.01 (1.03) | < 0.001 |
| Tau PET, mean (SD), SUVR |  |  |  |  | < 0.001 |
| - N missing | 946 | 179 | 77 | 1202 |  |
| - Mean (SD) | 1.08 (0.07) | 1.13 (0.08) | 1.23 (0.13) | 1.10 (0.09) |  |
| CDR Prog., No. (%) |  |  |  |  | < 0.001 |
| - CDR Non-progressor | 970 (77.2%) | 71 (28.1%) | 14 (11.8%) | 1055 (64.8%) |  |
| - CDR Progressor | 287 (22.8%) | 182 (71.9%) | 105 (88.2%) | 574 (35.2%) |  |

Individual and base LCMM (excluding Tau PET) mean trajectories are shown in Figure 1. Model selection criteria identified the model with two degrees of freedom and three latent classes, which we labeled as stable (77%), slow decliner (16%), and fast decliner (7%). The next best fitting model had three classes and three degrees of freedom (Δ BIC = 22.2, Δ ICL = 22.2). The best fitting Tau PET LCMM had two degrees of freedom and three latent classes (Supplementary Figure S3) and the next best fitting model had two classes and two degrees of freedom (Δ BIC = 376.4, Δ ICL = 336.9). The two models largely agreed on individual classifications. Only 7/427 (1.6%) individuals were reclassified from one model to the next, and none of those were fast decliners. Posterior class probabilities (Supplementary Figure S4) indicated good separation (mean posterior probability, 0.94), indicating model confidence in class assignment.

**Figure 1:**
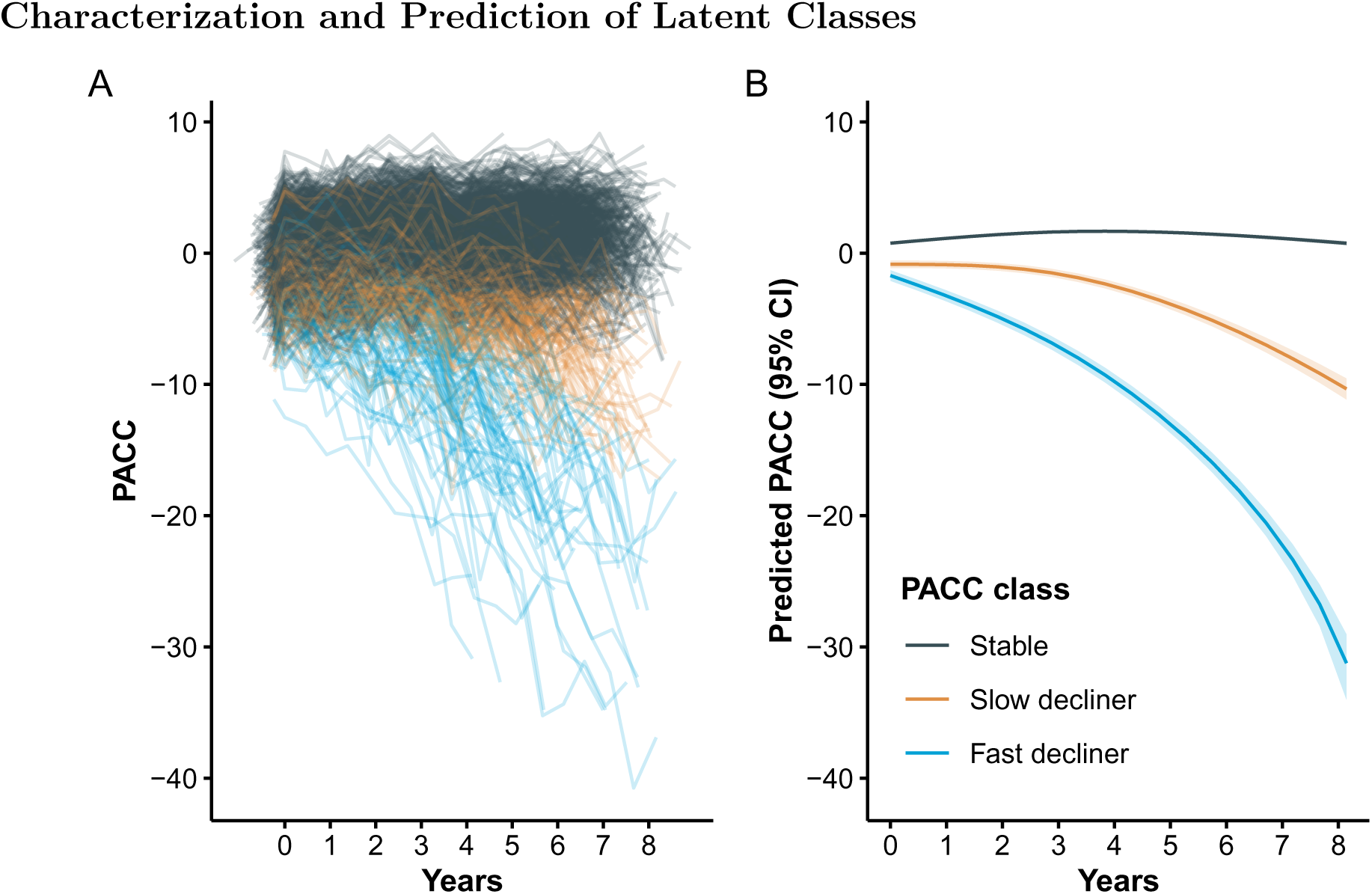
Individual and mean PACC trajectories by latent class. Left panel (A): Spaghetti plot of individual participant trajectories on the Preclinical Alzheimer Cognitive Composite (PACC), colored by latent class derived from the latent class mixed model (LCMM). Each line represents one participant’s observed scores over time. Right panel (B): Estimated mean PACC trajectories for each latent class, with shaded regions indicating 95% confidence intervals. Higher PACC scores indicate better cognitive performance.

The three classes are significantly separated starting immediately at baseline. From the base model, participants classified as fast decliners started with a mean -0.98 (95% confidence interval [CI] -1.44 to -0.52) PACC points and declined to a mean of -15.8 (95% CI -16.7 to -15.0) at six years. In contrast stable individuals started with a mean 0.52 (95% CI 0.37 to 0.67) PACC points and improved to a mean of 1.16 (95% CI 1.00 to 1.31) at six years. Slow decliners were intermediate between the other two classes starting with a mean -0.13 (95% CI -0.44 to 0.17) PACC points and declining to a mean -4.74 (95% CI -5.22 to -4.26).

Clinical relevance of the PACC latent classes was supported by differences in functional outcomes. The proportion of participants who were observed to progress on the Clinical Dementia Rating (CDR) Global score increased monotonically across latent classes, with the lowest rate among stable individuals (22.8%) and progressively higher rates among slow- (71.9%) and fast-decliners (88.2%) (Table 1). However, discordance between PACC latent class labels and observed CDR progression is common. While CDR Global scores greater than zero are possible without an indication of memory issues, this explains a very little of the discordance. Among CDR progressors, the proportion of individuals with a CDR Memory Box score of zero was 8.4% of PACC stable individuals, 5.5% of slow decliners, and 4.8% of fast decliners.

The distribution of continuous baseline variables by latent class is shown in Figure 2 and Supplementary Figure S1 summarizes standardized coefficients and 95% CIs from both components of the LCMM. Supplementary Figure S1 Panels A and C depicts odds ratios (ORs) for the class membership submodels from the base model and tau PET model. Continuous predictors are standardized so that ORs reflect the change in odds of belonging to the fast- or slow declining class compared with the stable class per one SD increase in each continuous baseline predictor.

**Figure 2:**
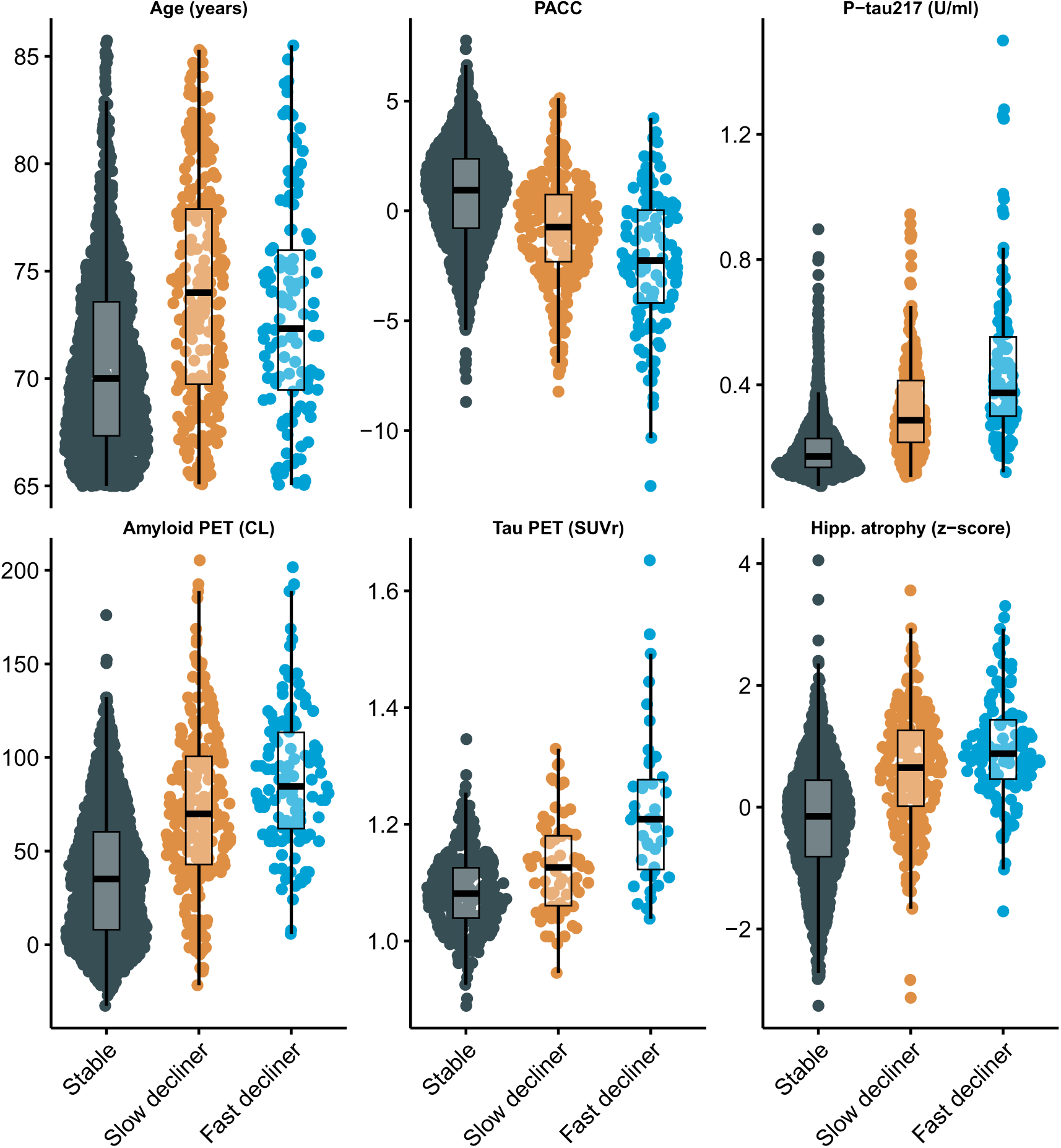
Distribution of baseline demographic and biomarker variables by latent class of cognitive decline. Sina plots show the distribution of selected baseline variables among latent classes identified by the latent class mixed model (LCMM) of Preclinical Alzheimer Cognitive Composite (PACC) trajectories: stable, slow decliner, and fast decliner. Variables include age, baseline PACC score, plasma phosphorylated tau 217 (P-tau217), amyloid positron emission tomography (PET) standardized uptake value ratio (SUVr), tau PET SUVr (subset with tau PET available), and hippocampal atrophy (volume normalized to intracranial volume). Each dot represents an individual participant; box plots represent quartiles. Higher PACC scores indicate better cognitive performance. **Abbreviations:** PET = positron emission tomography; P-tau217 = plasma phosphorylated tau 217; SUVr = standardized uptake value ratio; U/mL = arbitrary units proportional to assay signal.

**Figure 3:**
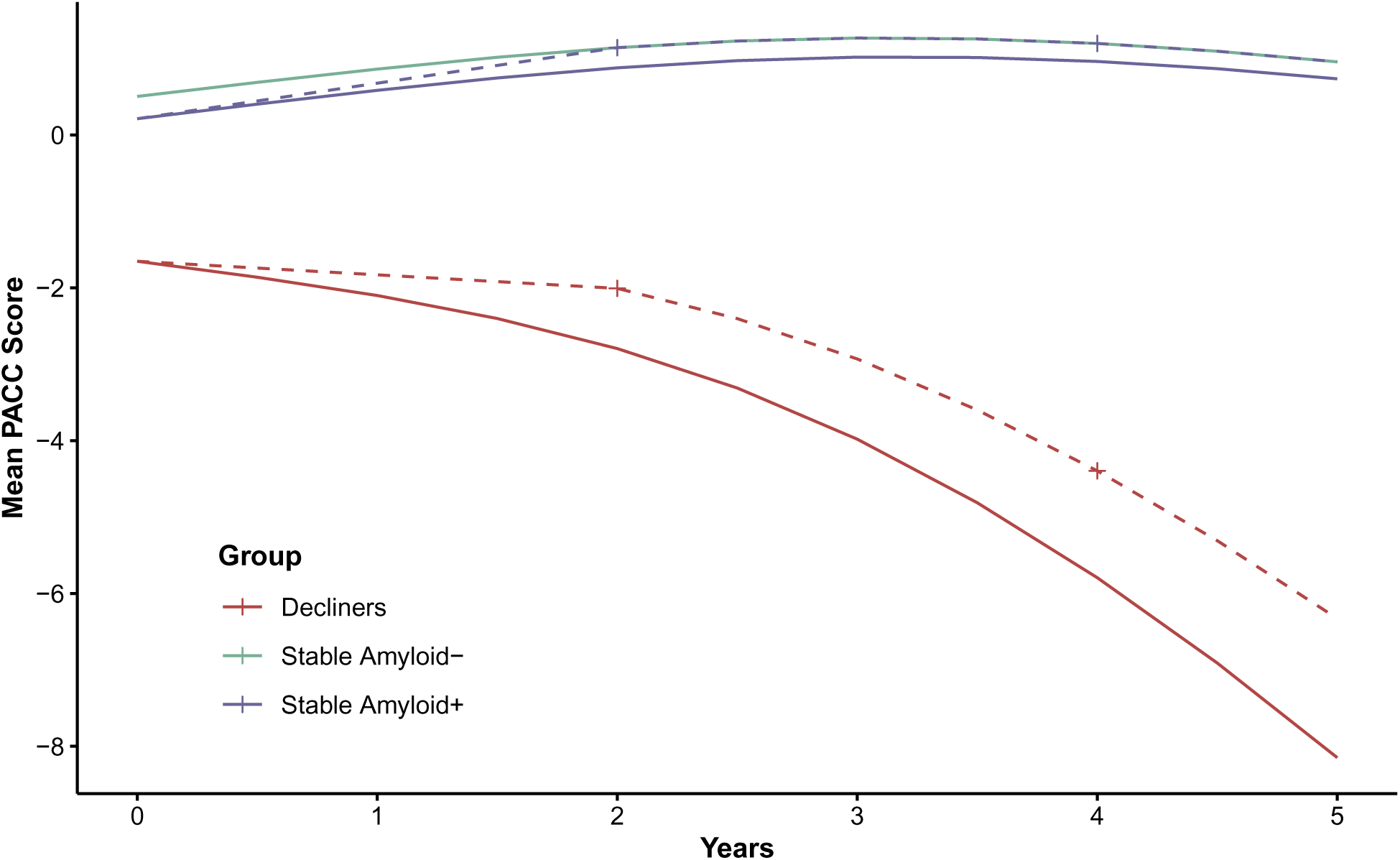
Trends for latent class-specific clinical trials. Mean trajectories were estimated from longitudinal models using natural cubic splines (two degrees of freedom) applied to Preclinical Alzheimer Cognitive Composite (PACC) scores for (a) two decliner classes, (b) amyloid-positive stable participants, and (c) a reference group of amyloid-negative stable individuals representing the maximum possible treatment benefit. For decliners, power calculations assume a treatment effect equal to 20% of the maximum possible benefit at 2 and 4 years. For amyloid-positive stable participants, power calculations assume a treatment effect equal to 100% of the maximum possible benefit at 2 and 4 years. Dashed lines represent projected mean PACC scores under treatment; crosses indicate time points used for power calculations.

From the base model (Panel A), higher plasma P-tau217 levels (OR 3.2, 95% CI 2.4 to 4.1), elevated amyloid PET centiloid (OR 1.4, 95% CI 1.0 to 1.9), and greater hippocampal atrophy scores (OR 3.9, 95% CI 2.7 to 5.6) were each associated with increased odds of belonging to faster-declining classes. Interestingly, P- tau217 and hippocampal atrophy had numerically stronger effects than amyloid PET. Also of interest, fast decliners were numerically younger (mean 73.1 years, SD 4.7), on average, than slow decliners (mean 74.1 years, SD 5.2) and increased age was associated with membership in the slow declining group (OR 1.4, 95% CI 1.1 to 1.7) but not the fast declining group (OR 0.93, 95% CI 0.69 to 1.2). Tau PET model results are largely similar, but confidence intervals are wider, likely due to smaller available sample (*N* = 427 vs *N* = 1, 629). Increased tau PET was associated with increased odds of slow- (OR = 1.8, 95% CI 1.2 to 2.8) and fast-declining groups (OR = 3.6, 95% CI 2.1 to 6.1).

Supplementary Figure S1 Panels B and D presents standardized coefficients from the longitudinal submodels, representing associations between each covariate and the PACC outcome. Higher PACC scores correspond to better cognitive performance and continuous variables are standardized and oriented so that positive coefficients indicate better performance. We see that females had increased risk of being fast decliners (OR 2.5, 95% CI 1.5 to 4.4), but had better PACC performance for a given class membership (0.30 PACC points, 95% CI 0.24 to 0.35). APOE *ε*4 carriage was associated with increased risk of both slow (OR 1.9, 95% CI 1.3 to 2.8) and fast (OR 2.9, 95% CI 1.6 to 5.3) declining class versus stable class, but negligible remaining effect on PACC. Education had no effect on class membership, but was associated with better PACC performance (0.12 PACC points per SD of education, 95% CI 0.095 to 0.15).

### Power Analysis for Latent Class-Specific Trials

Latent class-specific power analyses revealed marked differences in cognitive trajectories and variability across classes (see Supplementary Table S2). The amyloid-negative stable group had the highest mean (SD) PACC scores over time: +1.14 (2.11) at 2 years and +1.20 (2.25) at 4 years. We considered this the maximum possible treatment benefit. The stable amyloid-positive group had means of +0.88 (2.19) at 2 years and +0.96 (2.34) at 4 years. With 500 participants per arm, a trial in amyloid-positive stable individuals would have only 44% power at 2 years and 30% at 4 years to detect even 100% of the maximum benefit (0.26 PACC points at 2 years and 0.24 at 4 years). In contrast, a trial in amyloid-positive declining individuals would have 93% power at 2 years and 98% at 4 years to detect just 20% of the maximum benefit (0.79 PACC points at 2 years and 1.40 at 4 years).

These findings suggest that stable amyloid-positive individuals, although they may have the greatest potential long-term benefit from a disease-modifying drug, contribute little to the statistical power of a preclinical Alzheimer’s disease trial to detect an effect on the PACC. Instead, power is primarily driven by declining individuals, who represent a minority of the amyloid-positive population. Enrolling large numbers of stable individuals may dilute the overall treatment effect and reduce the trial’s ability to detect meaningful cognitive benefits. These results underscore the need to account for latent classes of decline when designing preclinical Alzheimer’s disease trials.

### Predictive Performance and Cross-Validation

The overall cross-validated accuracy was 0.80 (95% CI 0.79 to 0.82) for the base model and 0.756 (95% CI 0.73 to 0.78) for the tau PET model. However, this accuracy rate is inflated by the large number of stable individuals (1,257/1,629 = 77.2%), which are relatively easy for the model to identify. AUPRCs from the base model were 0.95, 0.41, and 0.49 for non-, slow-, and fast decliners versus the rest; and 0.70 for either slow- or fast decliner vs stable (Supplementary Figure S5). AUPRCs from the tau PET model were 0.92, 0.31, and 0.56 for non-, slow-, and fast decliners versus the rest; and 0.71 for either slow- or fast decliner vs stable (Supplementary Figure S6).

### Post-hoc Regression Tree

The optimally-tuned classification tree without tau PET achieved a mean balanced accuracy of 0.63 (standard error 0.014) in 10-fold cross-validation (Supplementary Figure S2). Variable importance analysis identified baseline P-tau217 (56.2%), baseline PACC (15.5%), amyloid PET (13.1%), and hippocampal atrophy (10.0%) as most important; though amyloid PET was not included in the tree, likely due to its correlation with P-tau217. The tree structure revealed a hierarchical decision process with participants with baseline P-tau217 < 0.29 were classified as stable individuals, though this miss-classifies *N* = 155 decliners. Those with P-tau217 ≥ 0.29 are further parsed by hippocampal atrophy, PACC, and P-tau217. Only one leaf contains a majority of fast decliners, and it contains only 1% of the initial sample. While the tree demonstrates the relative importance and utility of the predictors, it also demonstrates a high degree of miss-classification (31.4%). Cross-validated balanced accuracy is improved by the addition of tau PET (mean 0.70, standard error 0.011; Supplementary Figure S7). Tau PET has the second largest variable importance (22.5%), between P-tau217 (29.8%) and hippocampal atrophy (17.4%). The tree with tau PET results in a larger proportion leaves which are majority fast decliners (8.5% versus 1% without tau PET).

### Longitudinal Biomarker Progression Across Latent Classes

To further characterize biological differences among the latent trajectory groups, we compared longitudinal changes in amyloid PET, plasma P-tau217, tau PET (medial temporal and neocortical), and hippocampal volume across the four classes (Figure 4). As expected, slow and fast decliners showed the most rapid biomarker progression across modalities, consistent with their steeper cognitive decline. In contrast, A – stable individuals demonstrated minimal change over time in all biomarkers, supporting their classification as biologically and clinically stable. Notably, A + stable individuals, despite showing PACC trajectories that closely resembled A – stable individuals, exhibited clear evidence of biomarker progression, including increasing amyloid burden, rising plasma and PET tau signals, and accelerating hippocampal atrophy. These findings suggest that many A + stable individuals may represent an earlier stage of preclinical disease and could transition into declining classes with extended follow-up, highlighting the temporal dissociation between biomarker progression and short-term cognitive change.

**Figure 4:**
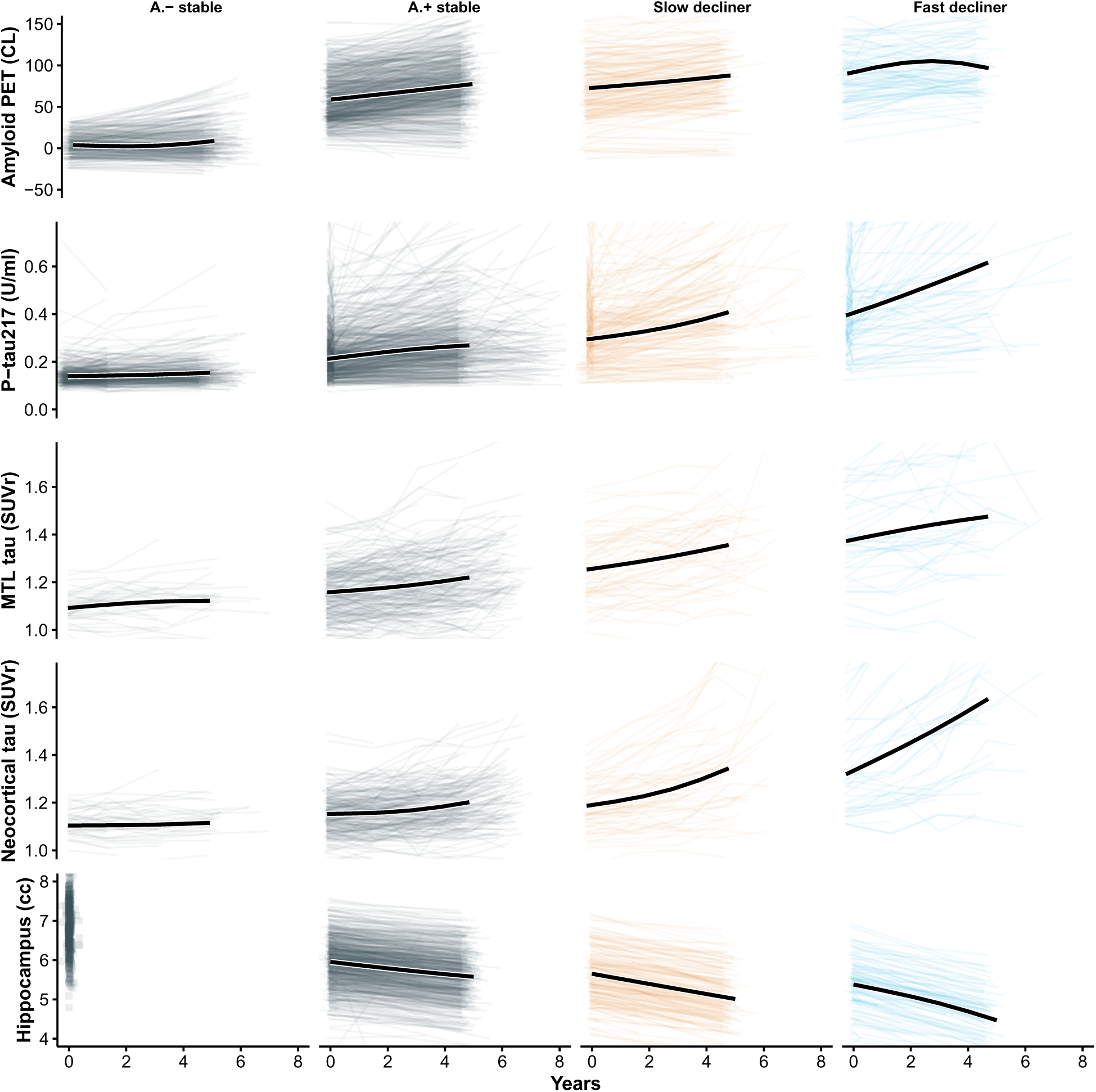
Longitudinal biomarker trajectories by PACC latent class. Each panel displays individual biomarker trajectories (light lines) and the estimated mean trajectory (bold black line) from a linear mixed-effects model with random slopes and natural cubic spline fixed effects (2 degrees of freedom). Columns correspond to A – stable, A + stable, slow decliner, and fast decliner. Rows represent five longitudinal biomarkers: amyloid PET (Centiloids), plasma P-tau217 (U/mL), medial temporal lobe (MTL) tau PET SUVR, neocortical tau PET SUVR, and hippocampal volume. While A – and A + stable individuals demonstrate similarly stable cognitive trajectories, the A + stable individuals exhibit clear biomarker progression across amyloid, tau, and neurodegeneration measures, suggesting that many may be in an earlier stage of preclinical disease and could transition into declining classes with longer follow-up.

## Discussion

In this latent class mixed-model analysis of cognitively unimpaired older adults, we identified distinct trajectories of PACC performance and evaluated the extent to which plasma and imaging biomarkers prospectively distinguished individuals who remained cognitively stable from those who showed slow or rapid decline. Several findings highlight challenges for prognostic modeling in the preclinical stage of Alzheimer disease and have direct implications for secondary prevention trial design.

### Available Predictors Only Partially Discriminate Future Decline

Although plasma P-tau217 and amyloid PET were each associated with latent class membership, neither biomarker alone, or in combination, achieved high accuracy in predicting of cognitive decline. A key observation was the high frequency of cognitive stability among amyloid-positive participants. As shown in Table 1, 776 of 1,110 amyloid-positive A4 participants (69.9%) were classified as stable individuals, indicating that substantial amyloid burden does not necessarily translate into measurable cognitive change over the timescales typical of prevention trials. However, this is consistent with a pre-symptomatic phase of disease believed to last as long as 15 years.

Compared to the 30.1% of amyloid-positive individuals classified as decliners, a small minority of amyloid-negative LEARN participants were classified as decliners. Thirty-eight of 519 (7.3%) amyloid-negative individuals were classified as slow or fast decliners, suggesting that some causes of decline may reflect non–AD processes, early AD pathology below PET detection thresholds at baseline that accumulated over the 5 year period, or cognitive measurement variability. These findings reinforce that amyloid positivity, while necessary for defining the AD pathophysiologic continuum, is an insufficient standalone predictor of short- to intermediate-term cognitive decline.

### Relative Contributions of Plasma P-tau217 and Tau PET

Plasma P-tau217 demonstrated stronger prognostic associations than amyloid PET, consistent with biomarker models positioning tau abnormalities closer to symptom onset. This observation aligns with prior reports, showing improved prognostic accuracy with plasma tau markers.^5^ However, classification tree analyses indicated that no single P-tau217 threshold meaningfully enriched for decliners without simultaneously excluding many true decliners, underscoring the limitations of threshold-based enrichment strategies.

Tau PET provided incremental predictive value beyond plasma P-tau217 and amyloid PET, although gains were modest. Because tau PET captures later-stage pathologic changes, these results are biologically plausible. Nevertheless, the logistical and financial limitations of tau PET constrain its feasibility as a routine screening tool in large-scale prevention trials.

### Implications for Secondary Prevention Trial Design

#### Participant Selection

Because most amyloid-positive individuals remain cognitively stable, eligibility based solely on amyloid positivity will enroll many stable participants, reducing power for cognitive endpoints.

Plasma P-tau217 may improve enrichment but cannot reliably isolate decliners. More complex multimodal strategies or repeated biomarker assessments may be needed for adequate prognostic discrimination.

#### Designing Trials for Heterogeneous Cohorts

Traditional power calculations assume a homogeneous population and uniform treatment effect size, overlooking substantial heterogeneity in cognitive trajectories. Fast decliners may show larger measurable effects but often represent individuals further along the disease continuum and exhibit greater variability, while stable individuals pose a different challenge. Their stability limits detectable cognitive benefit even if biological effects occur, requiring treatments to produce gains beyond natural improvement. Our analysis shows that variability across classes directly influences minimum detectable effect sizes, meaning that single-effect-size assumptions risk underpowered studies or inefficient resource allocation. More realistic simulation-based approaches that incorporate class-specific trajectories and differential treatment effects can improve trial design by informing enrichment strategies, optimizing sample sizes, and aligning outcomes with subgroup trajectories.

#### Added Value of Tau PET

Tau PET provided additional predictive information but its incremental value must be weighed against feasibility. Tau-PET-based staging may help identify individuals closer to clinical transition, though widespread implementation in prevention trials remains challenging.

#### Outcome Selection

Limited cognitive change in stable individuals raises questions about the sensitivity of traditional cognitive outcomes at this early stage. Digital measures of learning curves and biological endpoints such as plasma or tau PET may offer more responsive markers of treatment effect and serve as secondary or exploratory outcomes. Given evidence that tau PET correlates more strongly with cognitive decline than amyloid biomarkers, longitudinal tau PET or plasma P-tau217 may be useful for monitoring treatment response or detecting early trajectory divergence. Future work applying latent class or trajectory models to biomarker data may clarify whether biomarker-defined progressors can be identified earlier or more reliably than cognitive progressors.

#### Early Intervention

Anti-amyloid therapies show greater benefit in symptomatic individuals with less tau pathology, supporting the theory that earlier intervention yields greater benefit. However, if presymptomatic decline unfolds gradually over a decade or more, trials starting early may struggle to demonstrate cognitive benefit within feasible windows. Refining and validating sensitive digital and biomarker-based endpoints will be essential for advancing preclinical therapy development. Reliable measures of target engagement and disease progression could improve enrichment, increase power, and enable detection of treatment effects even among cognitively stable participants.

### Overall Interpretation

In summary, data-driven cognitive trajectories among initially unimpaired older adults exhibited substantial heterogeneity, and widely used biomarkers, including plasma P-tau217 and amyloid PET, were only partially effective in predicting who would decline over the time frame of a prevention trial. These limitations highlight the need for improved prognostic tools, potentially incorporating longitudinal biomarker dynamics, multimodal risk models, or digital assessments. Until then, secondary prevention trials must account for substantial heterogeneity in cognitive trajectories when planning enrollment, power calculations, trial simulations, and analytic strategies.

### Strengths and Limitations

This study has several strengths. We leveraged a large, deeply phenotyped cohort of cognitively unimpaired older adults with longitudinal cognitive assessments and harmonized biomarker data, using the A4LEARN R package to standardize and reproducibly analyze data across the A4 and LEARN cohorts. The latent class mixed-model approach allowed us to identify data-driven cognitive trajectories rather than relying on predefined thresholds or single-outcome definitions of decline. The inclusion of plasma P-tau217, amyloid PET, and, within a subset, tau PET enabled evaluation of multiple biomarkers spanning distinct phases of Alzheimer disease pathophysiology.

This study also has limitations. First, although latent class methods improve characterization of heterogeneity, class assignments are probabilistic and sensitive to model specification. Second, follow-up duration, while substantial, may still be insufficient to capture long-term cognitive change among stable individuals. Third, tau PET was available only in a subset, limiting statistical power for comparisons involving this biomarker and potentially reducing generalizability. Fourth, although we examined a broad panel of covariates, unmeasured factors, including comorbidities, lifestyle variables, and scanner-related differences, may influence both biomarker levels and cognitive outcomes. Finally, this is highly selected sample of individuals who were either eligible for the A4 study, or ineligible owing only to low amyloid PET signal, and may not fully represent community-dwelling older adults.

## Conclusions

Among cognitively unimpaired older adults, cognitive trajectories showed marked heterogeneity, and widely used biomarkers, including plasma P-tau217 and amyloid PET, were only partially effective in distinguishing who would decline. Tau PET provided modest additional predictive value but remains impractical for widespread enrichment. These findings highlight the challenge of prospectively identifying decliners in preclinical Alzheimer’s disease and underscore the need for improved multimodal prognostic tools. Secondary prevention trials should account for substantial heterogeneity in cognitive trajectories when developing enrollment strategies, powering cognitive endpoints, and interpreting treatment effects.

## Data Availability

All data used in the present study are available online at: https://www.a4studydata.org

## Acknowledgements

The authors would like to thank the A4 and LEARN Study Teams and site principal investigators and staff. Special gratitude to the A4 and LEARN participants and their study partners, without whom these studies would not be possible.

## Supplementary Material

**Supplementary Figure S1:**
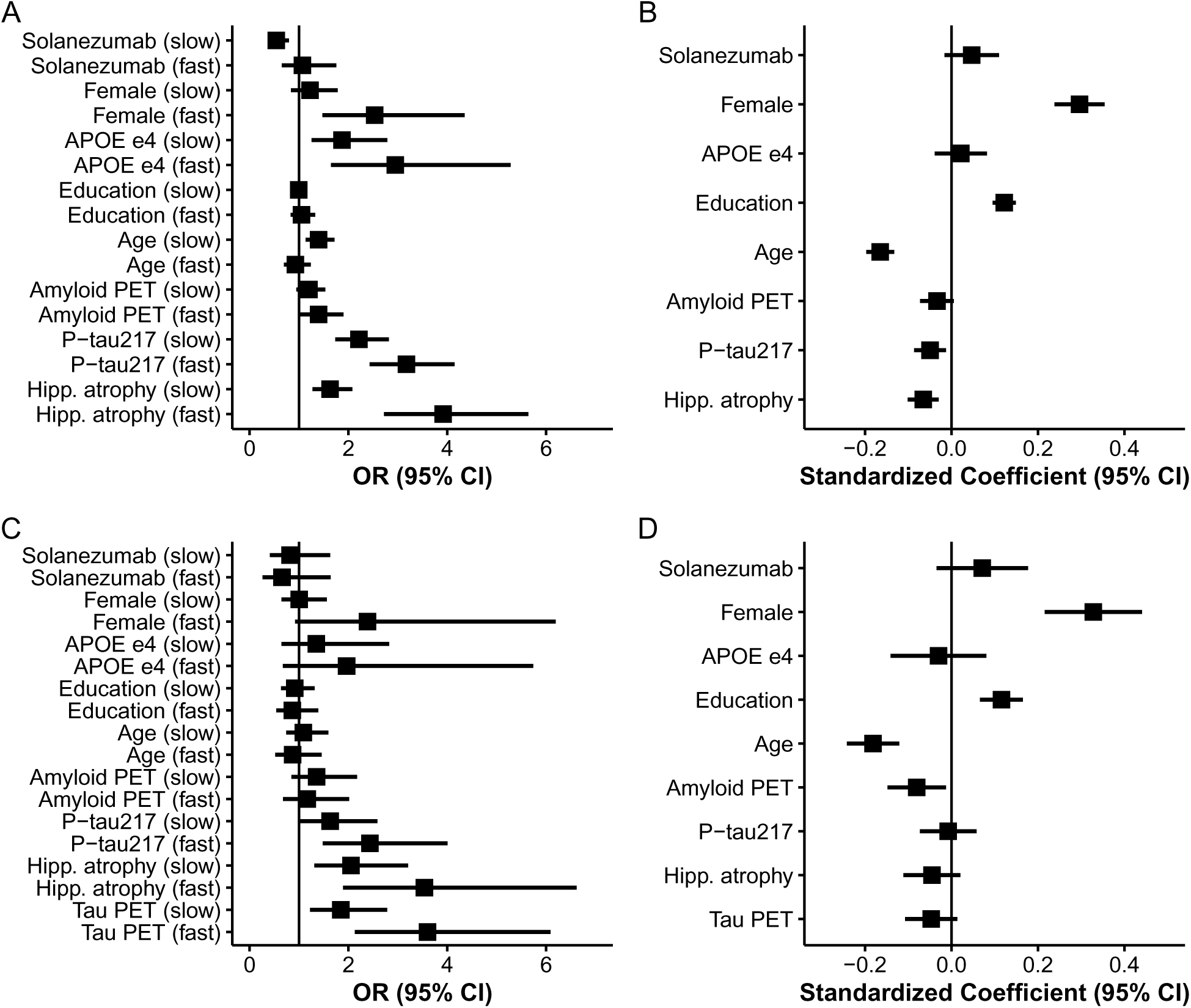
Association of baseline predictors with latent class membership and longitudinal cognitive decline in models with and without tau PET. Panels A and B display results from the latent class mixed model (LCMM) excluding tau PET; panels C and D show corresponding results from the model including tau PET as an additional predictor. (A, C) Odds ratios (95% CIs) from the LCMM class-membership submodel represent the relative likelihood of belonging to the slow- or fast-declining classes relative to stable class. (B, D) Standardized coefficients (95% CIs) from the LCMM longitudinal submodel show associations between baseline predictors and rate of change in the Preclinical Alzheimer Cognitive Composite (PACC). All continuous predictors and the PACC outcome were standardized (z-scored) before analysis to enable comparison of effect magnitudes. Higher PACC scores indicate better cognitive performance; thus, larger positive coefficients correspond to slower decline or better preservation of cognition over time.

**Supplementary Figure S2:**
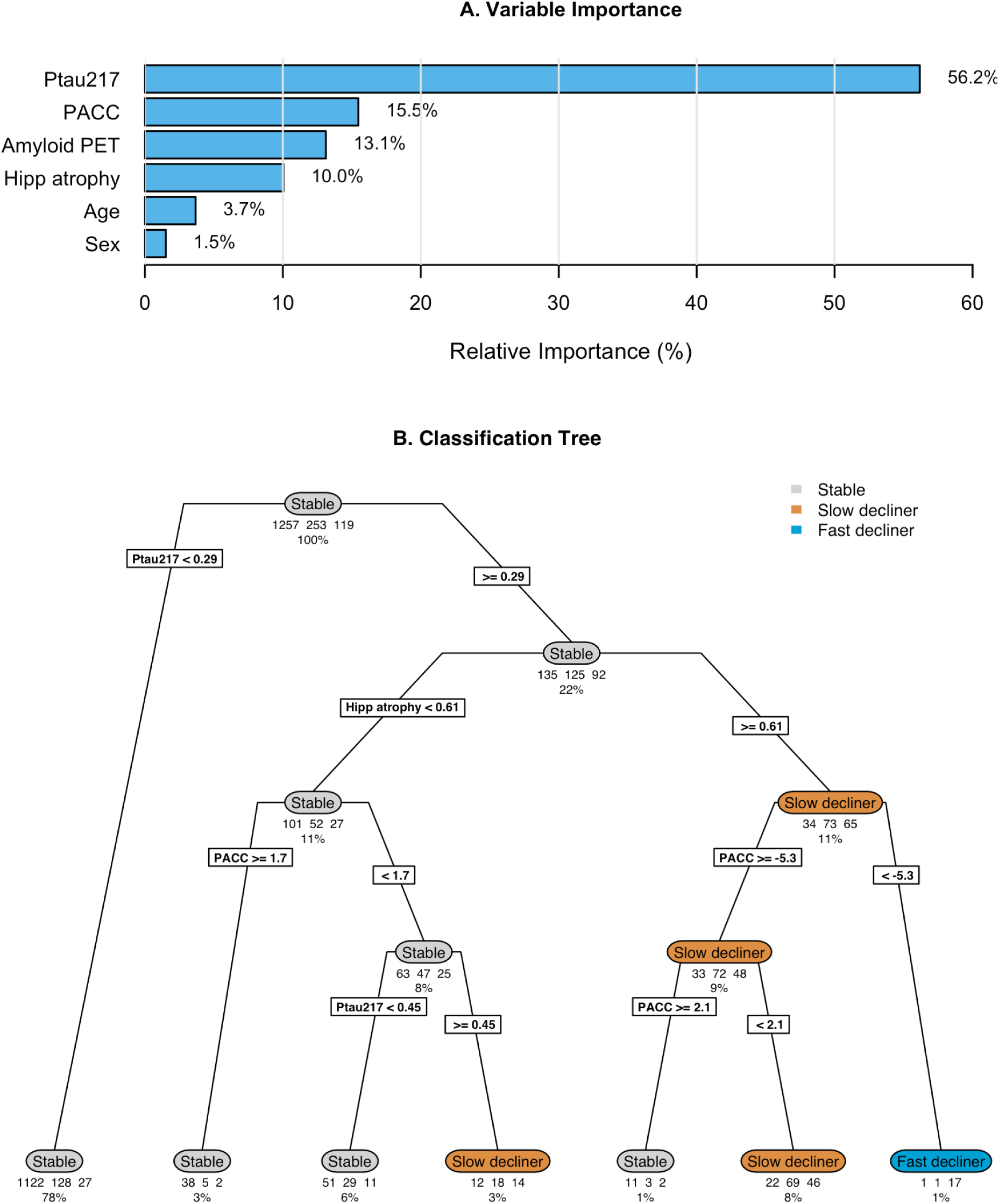
Classification tree analysis for latent class assignment from Base Model. Panel (A) shows relative importance of baseline predictors for class separation. Panel (B) shows classification tree showing binary decision rules for assigning subjects to latent classes based on predictors. Terminal nodes show predicted class, number of subjects, and percentage of total sample. The optimal tree complexity was determined using 10-fold cross-validation with the 1-standard error rule, resulting in a tree with 6 splits (mean balanced accuracy = 0.63, standard error 0.007)

**Supplementary Table S1:** Baseline Demographic, Clinical, and Biomarker Characteristics by Latent Class of Cognitive Decline with Tau PET. Baseline characteristics of participants classified by the latent class mixed model (LCMM) of Preclinical Alzheimer Cognitive Composite (PACC) trajectories. Classes represent distinct longitudinal cognitive patterns: stable, slow decliner, and fast decliner. Continuous variables are presented as mean (SD); categorical variables as No. (%). **Abbreviations:** APOE = apolipoprotein E; PET = positron emission tomography; P-tau217 = plasma phosphorylated tau 217; SUVr = standardized uptake value ratio; U/mL = arbitrary units proportional to assay signal; Am. = American; PI = Pacific Islander; CDR = Clinical Dementia Rating; Prog. = Progressor. **Footnotes:** Amyloid PET and tau PET SUVr values represent mean cortical uptake relative to cerebellar reference region. Hippocampal atrophy are residualized for intracranial volume and z-scored. Education reported in years of formal schooling. CDR Progressors were observed to have a CDR Global score greater than zero at two consecutive visits, or their last visit.

|  | Stable<br>(N=312) | Slow decliner<br>(N=73) | Fast decliner<br>(N=42) | Total<br>(N=427) | p value |
| --- | --- | --- | --- | --- | --- |
| Group |  |  |  |  | 0.004 |
| - LEARN | 52 (16.7%) | 3 (4.1%) | 0 (0.0%) | 55 (12.9%) |  |
| - Placebo | 131 (42.0%) | 34 (46.6%) | 23 (54.8%) | 188 (44.0%) |  |
| - Solanezumab | 129 (41.3%) | 36 (49.3%) | 19 (45.2%) | 184 (43.1%) |  |
| Age, mean (SD), y | 71.15 (4.51) | 73.74 (4.83) | 72.80 (5.65) | 71.76 (4.78) | < 0.001 |
| Sex, No. (%) |  |  |  |  | 0.009 |
| - Male | 126 (40.4%) | 40 (54.8%) | 11 (26.2%) | 177 (41.5%) |  |
| - Female | 186 (59.6%) | 33 (45.2%) | 31 (73.8%) | 250 (58.5%) |  |
| Race, No. (%) |  |  |  |  | 0.426 |
| - N missing | 3 | 0 | 1 | 4 |  |
| - Am. Indian or Alaska Native | 1 (0.3%) | 0 (0.0%) | 0 (0.0%) | 1 (0.2%) |  |
| - Asian | 12 (3.9%) | 2 (2.7%) | 4 (9.8%) | 18 (4.3%) |  |
| - Native Hawaiian or Other PI | 0 (0.0%) | 0 (0.0%) | 0 (0.0%) | 0 (0.0%) |  |
| - Black or African Am. | 7 (2.3%) | 4 (5.5%) | 0 (0.0%) | 11 (2.6%) |  |
| - White | 287 (92.9%) | 67 (91.8%) | 37 (90.2%) | 391 (92.4%) |  |
| - Unknown or Not Reported | 2 (0.6%) | 0 (0.0%) | 0 (0.0%) | 2 (0.5%) |  |
| Ethnicity, No. (%) |  |  |  |  | 0.842 |
| - Hispanic or Latino | 5 (1.6%) | 1 (1.4%) | 1 (2.4%) | 7 (1.6%) |  |
| - Not Hispanic or Latino | 302 (96.8%) | 70 (95.9%) | 41 (97.6%) | 413 (96.7%) |  |
| - Unknown or Not reported | 5 (1.6%) | 2 (2.7%) | 0 (0.0%) | 7 (1.6%) |  |
| Education, mean (SD), y | 16.24 (2.89) | 16.16 (2.66) | 16.05 (2.70) | 16.21 (2.83) | 0.904 |
| APOE 4, No. (%) |  |  |  |  | < 0.001 |
| - non-carrier | 166 (53.2%) | 22 (30.1%) | 11 (26.2%) | 199 (46.6%) |  |
| - carrier | 146 (46.8%) | 51 (69.9%) | 31 (73.8%) | 228 (53.4%) |  |
| PACC, mean (SD) | 0.51 (2.48) | -1.20 (2.57) | -2.57 (3.20) | -0.09 (2.77) | < 0.001 |
| Amyloid PET, mean (SD), CL | 47.04 (30.54) | 77.33 (38.73) | 86.23 (34.78) | 56.07 (35.74) | < 0.001 |
| P-tau217, mean (SD), U/ml | 0.22 (0.10) | 0.33 (0.17) | 0.43 (0.20) | 0.26 (0.15) | < 0.001 |
| Hipp. atrophy, mean (SD), z-score | 0.03 (0.83) | 0.80 (0.85) | 0.96 (0.79) | 0.25 (0.91) | < 0.001 |
| Tau PET, mean (SD), SUVR | 1.08 (0.07) | 1.13 (0.08) | 1.23 (0.13) | 1.10 (0.09) | < 0.001 |
| CDR Prog., No. (%) |  |  |  |  | < 0.001 |
| - CDR Non-progressor | 217 (69.6%) | 18 (24.7%) | 2 (4.8%) | 237 (55.5%) |  |
| - CDR Progressor | 95 (30.4%) | 55 (75.3%) | 40 (95.2%) | 190 (44.5%) |  |

**Supplementary Figure S3:**
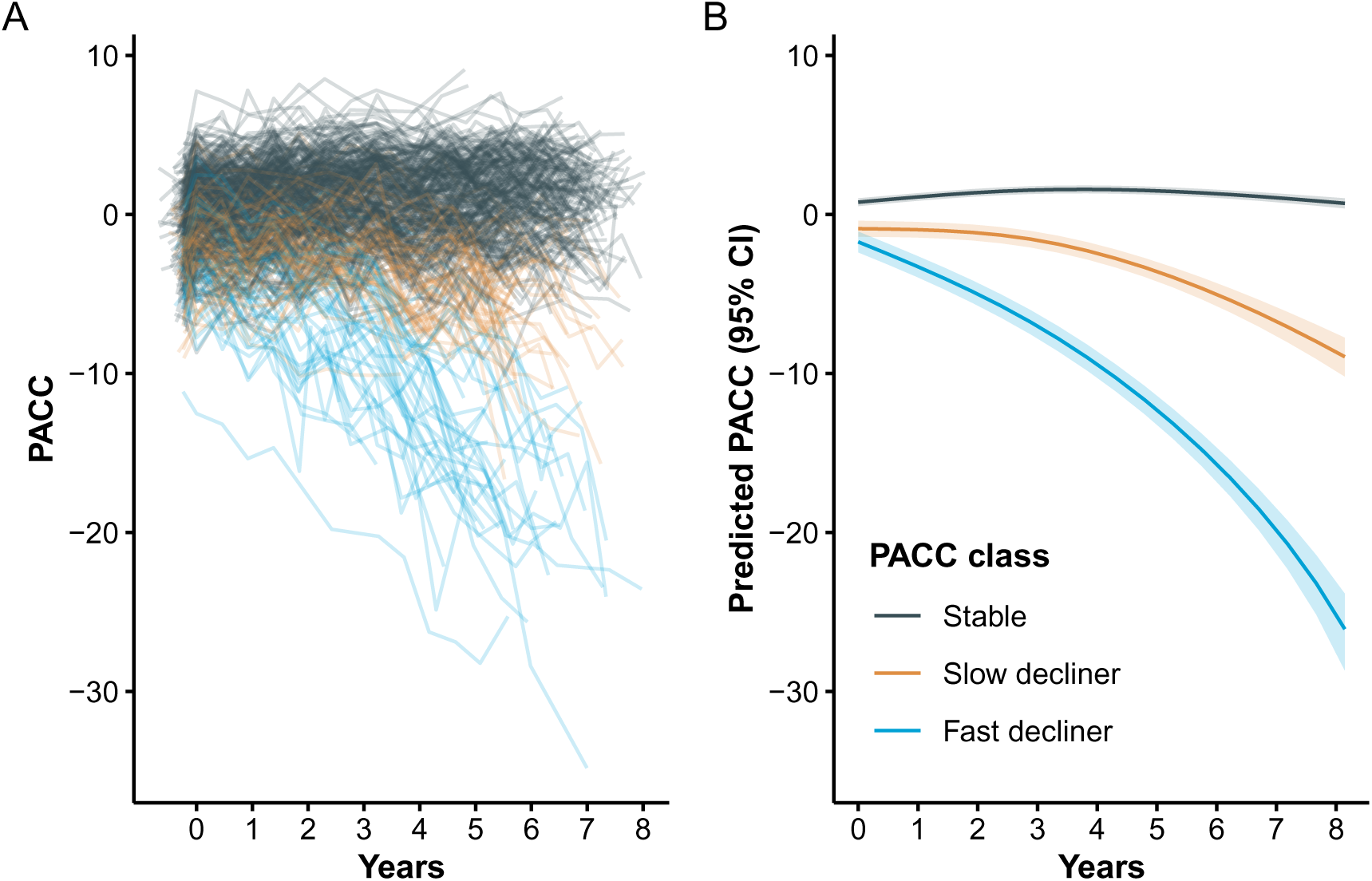
Individual and Mean PACC Trajectories by Latent Class Model with Tau PET. Left panel (A): Spaghetti plot of individual participant trajectories on the Preclinical Alzheimer Cognitive Composite (PACC), colored by latent class derived from the latent class mixed model (LCMM) which included tau PET as a predictor. Each line represents one participant’s observed scores over time. Right panel (B): Estimated mean PACC trajectories for each latent class, with shaded regions indicating 95% confidence intervals. Higher PACC scores indicate better cognitive performance. **Abbreviations:** PET = positron emission tomography.

**Supplementary Figure S4:**
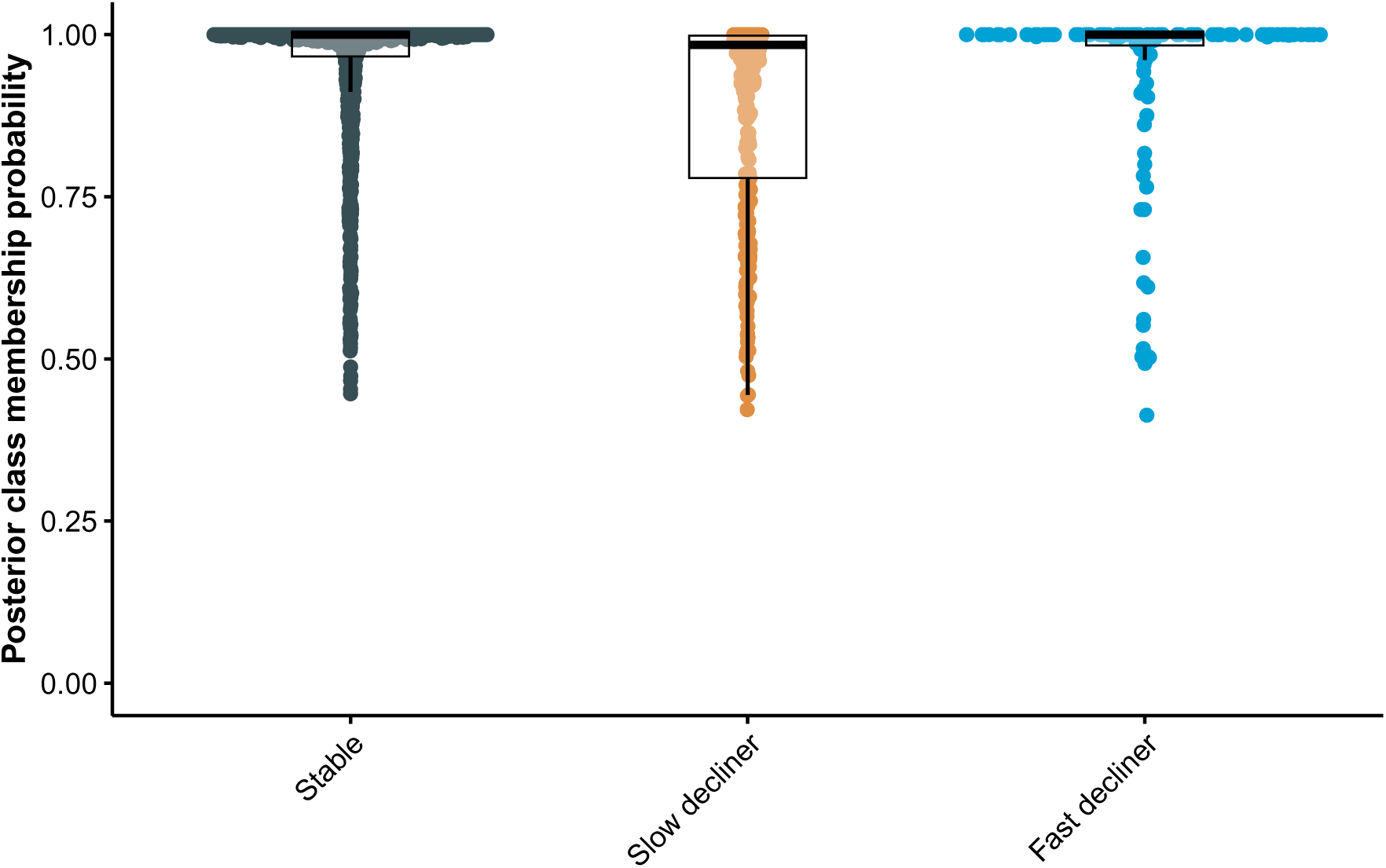
Distribution of posterior class membership probabilities by assigned latent class, indicating model confidence in class assignment. This plot shows the degree of certainty in class assignments, a good indicator of model reliability. Mean posterior probabilities were {r}pp_sum[[1,2]], {r}pp_sum[[2,2]], and {r}pp_sum[[3,2]] for non-, slow-, and fast decliners. With most participants having high posterior probabilities (> 0.8) for their assigned class, class separation is strong and predictive discrimination is good.

**Supplementary Figure S5:**
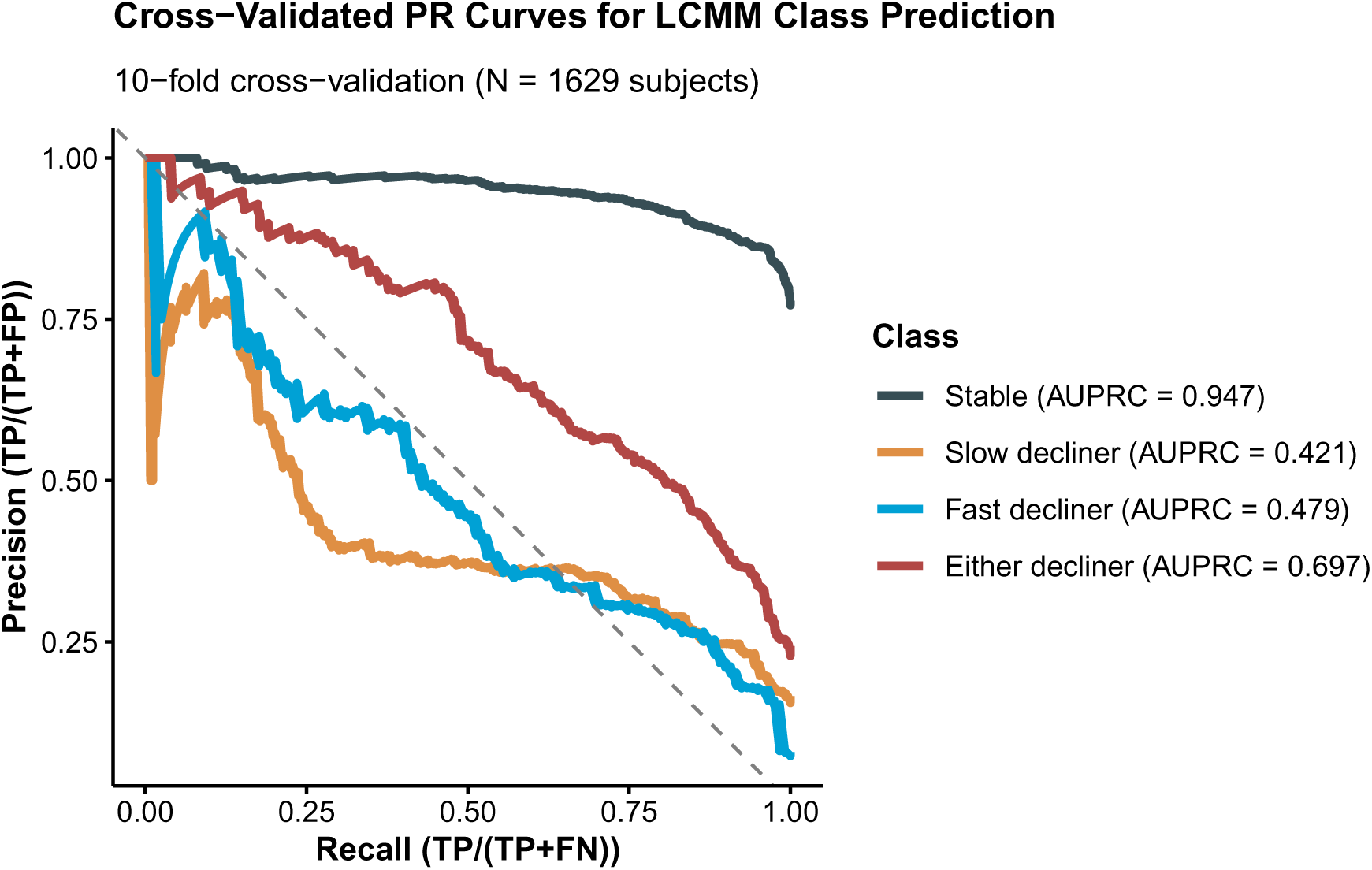
Cross-validated Precision-Recall Curves for Latent Class Membership Model Without Tau PET. Precision-recall curves summarizing 10-fold cross-validated classification performance of the latent class membership submodel based on baseline demographics and biomarkers, excluding tau PET. Curves depict the ability to distinguish the given class from the other two. The area under the precision-recall curve (AUPRC) quantifies overall predictive performance, emphasizing recall (or sensitivity, TP/(TP+FN)) and precision (TP/(TP+FP)) for less prevalent classes. **Abbreviations:** AUPRC = area under the precision-recall curve; TP = true positive rate; FN = false negative rate; FP = false positive rate.

**Supplementary Figure S6:**
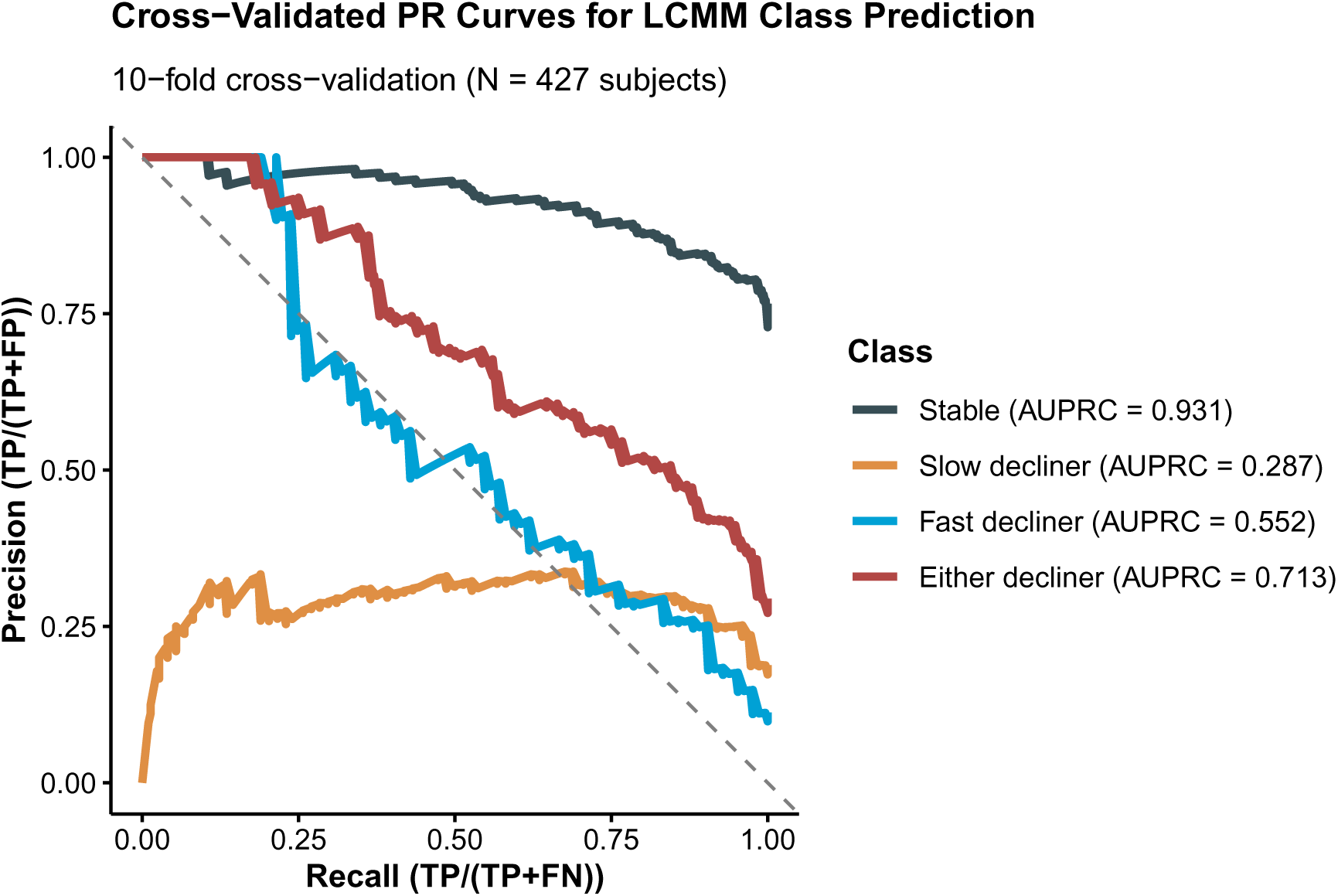
Cross-validated Precision-Recall Curves for Latent Class Membership Model With Tau PET. Precision-recall curves summarizing 10-fold cross-validated classification performance of the latent class membership submodel based on baseline demographics and biomarkers, excluding tau PET. Curves depict the ability to distinguish the given class from the other two. The area under the precision-recall curve (AUPRC) quantifies overall predictive performance, emphasizing recall (or sensitivity, TP/(TP+FN)) and precision (TP/(TP+FP)) for less prevalent classes. **Abbreviations:** AUPRC = area under the precision-recall curve; TP = true positive rate; FN = false negative rate; FP = false positive rate.

**Supplementary Figure S7:**
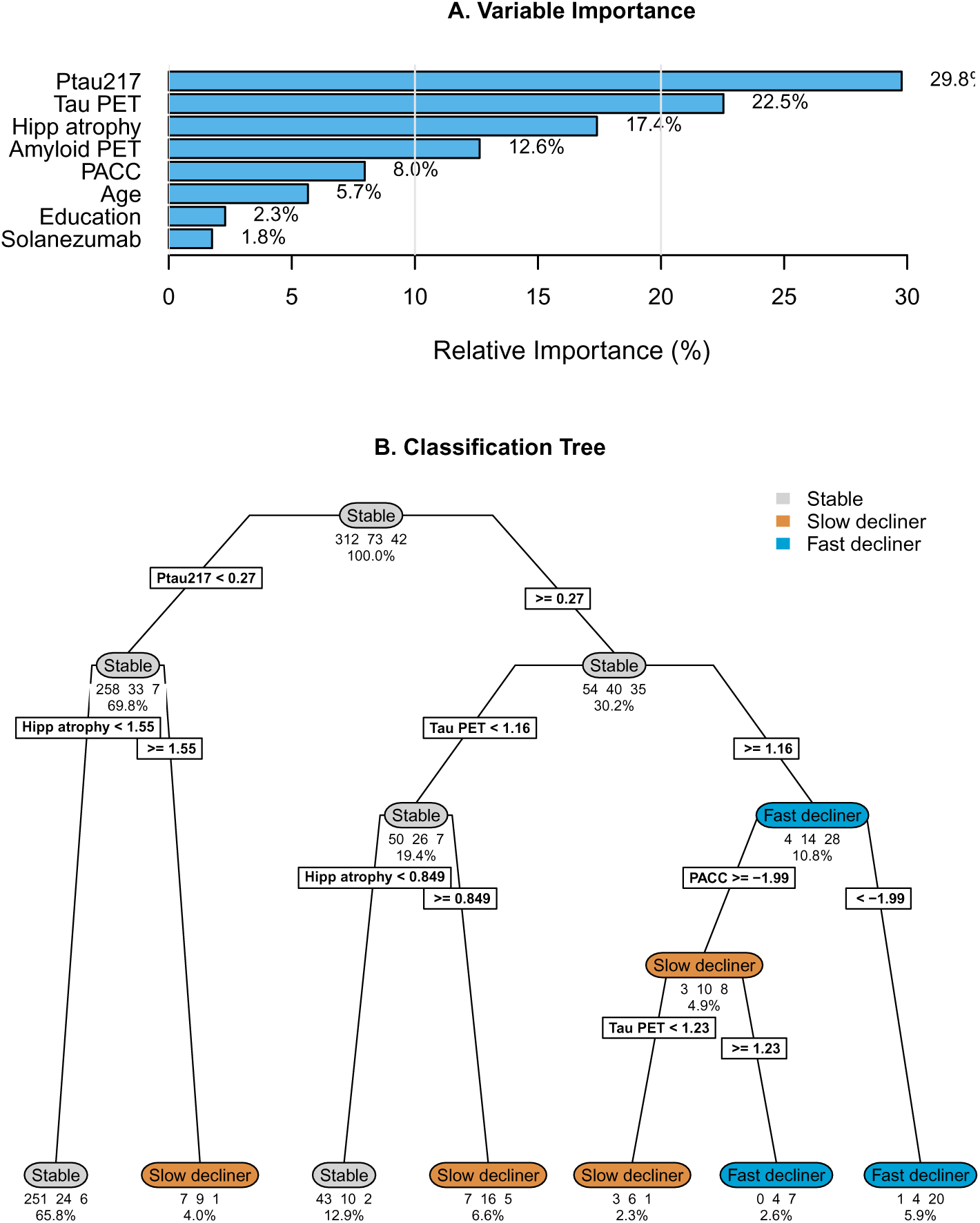
Classification tree analysis for latent class assignment from Tau PET Model. (A) Relative importance of baseline characteristics for class separation. (B) Classification tree showing binary decision rules for assigning subjects to latent classes based on baseline characteristics. Terminal nodes show predicted class, number of subjects, and percentage of total sample. The optimal tree complexity was determined using 10-fold cross-validation with the 1-standard error rule, resulting in a tree with 6 splits (mean balanced accuracy = 0.70, standard error 0.011).

**Supplementary Table S2:**
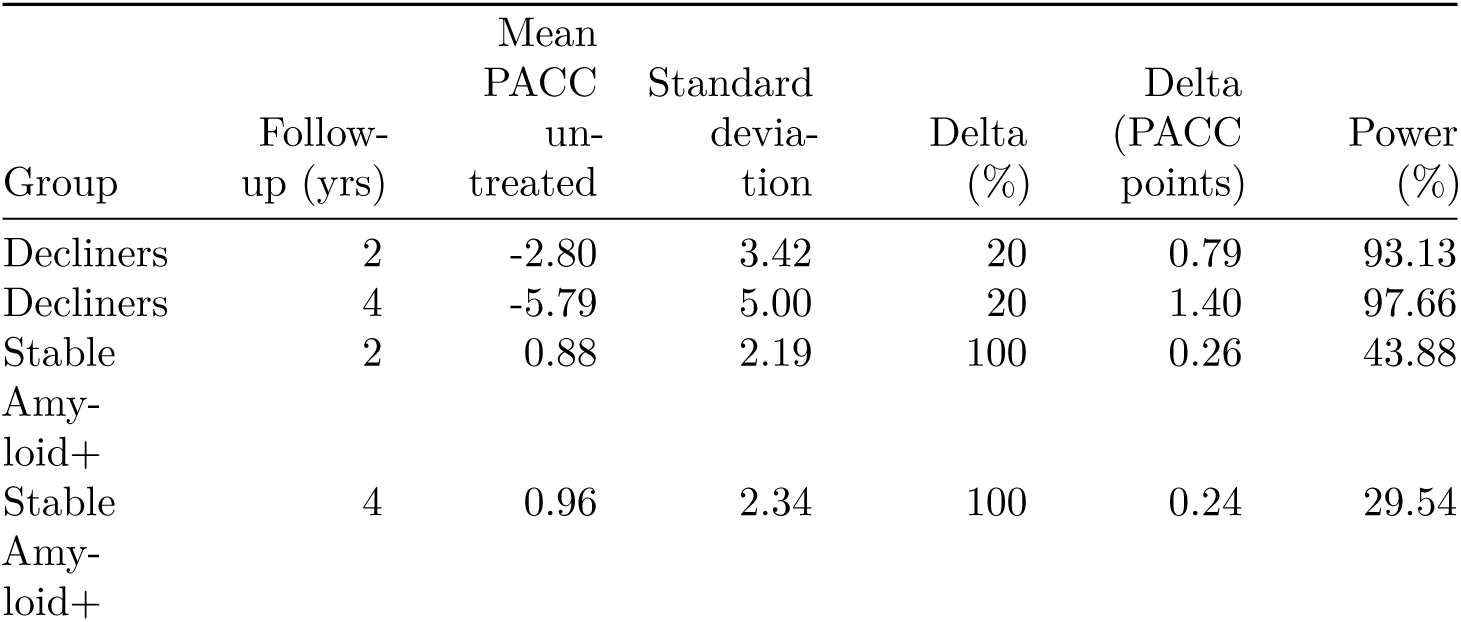
Power for latent class-specific clinical trials. Effect size estimates were derived from longitudinal models using natural cubic splines (two degrees of freedom) applied to Preclinical Alzheimer Cognitive Composite (PACC) scores among amyloid-positive A4 participants in (a) the stable class and (b) the two decliner classes. The table reports mean PACC and residual variance at 2 and 4 years, representing expected control group trajectories for two potential trial durations. Power was approximated using two-sample t-test calculations assuming 500 participants per group, with 10% attrition at 2 years and 20% at 4 years. Effect size is expressed both as absolute PACC change and as a percentage of the maximum possible improvement, defined as the mean PACC among stable amyloid-negative LEARN participants at the corresponding time point.

